# Cardiovascular Risk Factors Identified Among Homeless Adults in San Francisco: Recommendations for Evidence-Based Outreach Services From a Quality Improvement Initiative

**DOI:** 10.64898/2026.03.22.26349032

**Authors:** Sarah Jean Valliant, Ileana Jade Rodriguez, Angelina Ka Lee, Carole M. Kulik, Rianna Rose Punzalan, Logan Charles Holbrook, Raquel Juliette Tamayo, Roxanne Carlos Mendoza, Macarena Sofia Puig, Trevor Anderson, Yacoob Ebrahim Modan, Simran Athwal, Isaak Lugo, Melissa Hernandez-Monroy, Destiny-Elizabeth Auraya Silva-Castro, Michael Petrides, Nyssa Alvarado, Kelly Tang

**Author notes:** Corresponding Author: Sarah Jean Valliant.

## Abstract

**Objective:** This preliminary public health report assessed acute and chronic health burdens, focusing on cardiovascular health, among unsheltered individuals experiencing homelessness. It aims to guide medical referrals, deliver targeted health education, and prioritize services within a community-based nonprofit.

**Methods:** A field-based needs assessment used a structured questionnaire to evaluate acute and chronic health burdens. Clinical measures included blood pressure (BP), heart rate (HR), pain scores (normalized to 0–10), nicotine use, and diabetes prevalence. Of 72 initial responses, 59 BP, 65 HR, and 66 pain scores were usable. BP was classified per ACC/AHA (2017) guidelines [1], including Hypertensive Crisis. Nicotine and diabetes data from a secondary survey yielded 39 and 38 usable responses of 116. Ethical oversight ensured informed consent, participant capacity assessment, and emergency protocols. Data were analyzed descriptively.

**Results:** Participants were predominantly male (N = 53 of 72) with ages ranging from 24 to 70 years (Mean = 42.96; Median = 41; N = 70). The cohort was primarily White/Caucasian (N = 30) and Black/African American (N = 27). Cardiovascular assessments revealed substantial acute risk: 72.88% (N = 43 of 59) of BP readings were classified as Total High Blood Pressure, and 10.17% (N = 6 of 59) met criteria for Hypertensive Crisis or higher, including readings of 210/137 mmHg and 286/127 mmHg. Mean and median HR were both 96 bpm (N = 65). Chronic symptom burden was notable, with a mean pain score of 3.74 and 19.70% (N = 13) reported severe pain (7–10). Self-reported comorbidities included current smoking in 15.38% (N = 6 of 39) and a history of diabetes in 13.16% (N = 5 of 38).

**Conclusion:** Findings show a high prevalence of acute cardiovascular risk, particularly severe hypertension, among the unsheltered population. These results highlight the urgent need for improved outreach, targeted cardiovascular and primary care referrals, and follow-up screenings. Expanding health education on the effects of uncontrolled diabetes and smoking is recommended to reduce future cardiovascular events.

**Highlights:**

- Hypertensive Crisis prevalence: 10.17% (N = 6) of usable BP ratios (N = 59) met the criteria for Hypertensive Crisis or Higher (SYS ≥ 180 OR DIA ≥ 120).
- Overall prevalence of High Blood Pressure (Stage 1 or higher): 72.88% (N = 43) of usable BP ratios (N = 59), representing a pervasive, unmanaged health condition.
- Elevated Mean Cardiac Activity: The calculated Mean Heart Rate was 96 (N = 65 usable entries), with a Median Heart Rate of 96, suggesting chronic stress or underlying conditions.
- High Chronic Pain Burden: 19.70% (N = 13) of participants reported severe pain scores of 7-10 (N = 64 usable scores after transformation).
- Demographic Context: The cohort included 72 usable responses with a mean age of 42.96 years. Sex distribution records 55 males, 13 females, and 4 unknown/missing entries. Race distribution records 30 White/Caucasian, 27 Black/African American, 3 Hispanic/Latinx, 2 Native American/Indigenous American, six other/unspecified, and four unknown/missing entries.
- Nicotine Use Prevalence: 15.38% (N = 6) of participants reported nicotine use. All forms of nicotine products, including cigarettes and smokeless tobacco, were combined for analysis, while non-nicotine product substances were excluded.
- Diabetes Prevalence: 13.16% (N = 5) of participants self-reported a diagnosis of diabetes. All forms of diabetes were combined into a single variable for analysis.

## Introduction

### Background

The unsheltered and homeless community is globally recognized as a population facing extreme health disparities, often characterized by premature mortality and high rates of hypertension, congestive heart failure, myocardial infarctions, and other chronic diseases, with poor cardiovascular outcomes coupled with limited access to continuous medical care. [2,3] Nationally, individuals experiencing homelessness have mortality rates three to four times higher than the general population, with cardiovascular disease identified as one of the leading causes of death within this group. [4]

Despite growing literature on health disparities, few studies have quantified acute cardiovascular risk factors using field-based clinical measurements among unsheltered adults. Existing studies rely on self-reported or retrospective health measures, while field-based data can directly quantify cardiovascular risk factors. Field-based measurements will reflect the immediate needs of unsheltered adults, addressing a critical knowledge gap in public health surveillance, intervention planning, and resource allocation.

Cardiovascular disease prevalence among people experiencing homelessness has been found to affect these individuals with increased risks for mortality at disproportionate levels when compared to housed populations. Reasons for this disparity include heightened lifestyle risk factors such as stress, smoking, illicit drug use, and a lack of overall health maintenance, coupled with the difficulty of identifying or diagnosing cardiovascular disease in unsheltered adults and subsequently providing the appropriate interventions, treatment, and management of disease [5]. Poor management of cardiovascular disease can lead to devastating health outcomes such as acute cardiovascular events like myocardial infarctions (MI), cerebrovascular accidents (CVA), and even premature death. A report by the National Academies of Sciences, Engineering, and Medicine identified poor social health as a significant contributor to cardiovascular disease (CVD) risk, associated with a one-third increase in CVD incidence and a fourfold relative increase in the risk of mortality, hospitalization, and emergency department visits. [6]

When considering other risk factors that can lead to cardiovascular pathology, diabetes mellitus (DM) and smoking are two known factors that can cause vascular damage. Both DM and smoking tobacco products are known to increase risks for atherosclerosis, MI, stroke, and overall mortality [7].

Diabetes Mellitus (DM) is known to be a driving factor in cardiovascular disease due to the multi-factorial pathophysiology of DM. Hyperglycemia, the hallmark symptom of DM, has been found to accelerate atherosclerosis through microinjuries such as nephropathy, neuropathy, and retinopathy while creating macroinjuries that promote atherosclerosis, such as advanced glycation end-products, which ultimately modify lipids and proteins [8, 9]. Glycated LDL cholesterol is cleared less efficiently, promoting plaque formation [9].

The prevalence of CVD in adults who smoke cigarettes or tobacco products is projected to be at least 2-4 times more likely than in a healthy and non-smoking adult [10]. Studies suggest that smoking is the initiating and progressive factor of atherosclerotic heart disease and vascular injuries, as smoking has multiple biological pathways of damaging blood vessels, including endothelial dysfunction, inflammation, platelet activation, and enhancing thrombosis [10].

Previous studies do note that the risk for CVD increases synergistically when there is a combined history of DM and smoking-heavy smokers with DM are 50% more likely to develop CVD compared to a non-smoker with DM, and 20% more likely to develop CVD than a healthy and non-smoker adult [7]. While studies observing the synergistic rather than additive effect of smoking and DM on individuals experiencing homelessness are limited, it is crucial to understand that DM and smoking independently have negative impacts on cardiovascular health, and when both conditions are present, the implications for cardiovascular health increase.

Pain is often recognized as a subjective symptom, yet growing evidence indicates it is a risk factor for cardiovascular morbidity. Chronic pain and multisite pain have been identified as contributors to adverse cardiovascular outcomes [11], supported by recent findings that chronic and multisite pain were strongly associated with cardiovascular events of MI and CVA; incidents of MI occurring at a rate of 1.48 per 1000 person-years and CVA occurring at a rate of 0.86 per 1000 person-years [12].

### Objectives

This public health report was conducted exclusively as an Operational Needs Assessment (ONA) for Valliant Foundation, rather than as formal research determined by an independent ethics committee. The primary aim was to understand baseline health status, acute cardiovascular risk factors, and chronic symptom burden among unsheltered individuals to ethically and effectively inform the Foundation’s operational strategies. The assessment also highlights the elevated prevalence of co-occurring conditions, including nicotine use and diabetes, which substantially contribute to the overall health burden within this population. By systematically documenting these health risks, the findings provide critical insights for public health stakeholders, emphasizing the urgent need for targeted interventions, community-based education, and resource allocation to mitigate preventable morbidity and enhance health equity among unhoused communities.

#### The specific objectives of this assessment were to

##### 1. Quantify Acute Cardiovascular Risk

To assess and characterize the prevalence of acute, potentially life-threatening cardiovascular risk factors, specifically blood pressure and heart rate (BP, HR) [13] within the unsheltered population, providing evidence to guide the prioritization of medical referrals and the activation of the Emergency Action Protocol.

##### 2. Characterize Chronic Symptom Burden

To quantify the prevalence and severity of chronic pain within the cohort, using transformed pain scores, to prioritize resource allocation toward pain management and quality-of-life interventions, as the presence of severe pain can have negative impacts on cardiovascular status. [14]

##### 3. Establish a Demographic and Health Profile

To describe the basic demographic profile (age, gender, race/ethnicity) of the participants to ensure future outreach efforts, educational materials, and service designs are appropriately targeted and culturally competent.

##### 4. Identify Other Cardiovascular Risk Factors

To measure the possible impacts of smoking and diabetes on cardiovascular health, and to provide appropriate health education.

#### Rationale

To reduce health inequalities within the homeless community, outreach programs need to acquire a holistic understanding of the baseline health status, acute risk factors, and chronic health burden. This public health report provides a multi-metric health assessment conducted within a local homeless outreach initiative to produce data that will strengthen and guide future intervention and prevention strategies. A critical finding demonstrates that homeless adults have the highest rates of smoking, cocaine use, and suboptimal control of cardiovascular risk factors [4], highlighting their vulnerability to preventable health outcomes. Illustrating this foundational data enables prioritizing medical referrals, health education, and advocacy efforts to ensure resources are allocated to individuals facing the highest cardiovascular risk.

Drawing on data from 116 participant responses, this provided valuable data for a focused clinical profile based on five metrics: heart rate (HR), blood pressure (BP), pain scores, and self-reported smoking and diabetes status. BP and HR values are real-time indicators of life-threatening cardiovascular instability and physiological function, while pain scores serve as a subjective screening for experienced distress. Any responses of BP readings over 180/120 mmHg informed outreach staff of hypertensive-related emergencies and urgent intervention to prevent the risk of stroke or heart attack. HR provides real-time information on extreme or irregular heart rates, reflecting the body’s current physiological state. Suppose the HR value recording indicates an abnormal heart rate. In that case, it may indicate various health problems, including cardiovascular disease, stroke, and heart failure, and suggests immediate attention to stabilize abnormal measurements. [15]. Lastly, pain scores on a 1-10 scale indicate the level of urgent attention needed, even if vital signs appear exceptionally normal. The direct clinical readings were complemented by participants’ self-reporting of both smoking status and a history of diabetes. Smoking and diabetes are two factors that significantly increase cardiovascular risk and mortality within the homeless population. [16]. Collectively, these metrics inform volunteers on how to provide referrals, activate emergency response systems, and deliver education in a professional, time-efficient manner.

### Methodology

#### Ethical Oversight and Project Definition

This report summarizes data collected through a community needs assessment conducted by Valliant Foundation’s outreach program. The activity was strictly defined as an Operational Needs Assessment, not a research project, with the sole purpose of gathering descriptive data to guide non-profit operational decisions regarding resource prioritization, referrals, educational materials, and services.

Before commencement, the project underwent review by Valliant Foundation’s *Ethics Committee*, an independent oversight body authorized under the Foundation’s bylaws to review, approve, and enforce ethical standards for operational activities. The Committee determined that this project constitutes Non-Human Subjects Research (NHSR) under 45 CFR 46.102, as it does not aim to contribute to generalizable knowledge and was conducted solely to inform nonprofit operations and outreach programming (Reference No: V5671).

Inquiries regarding this determination may be directed to Valliant Foundation Ethics Committee for further clarification, interest, or guidance at

### Study Setting and Participants

The assessment was conducted in San Francisco, California, from November 2024 to September 2025 as part of regular outreach activities targeting the unsheltered community. Specific San Francisco communities served included the Inner Mission District, Tenderloin, and the Embarcadero. Data were collected only during daylight hours and in good weather to ensure environmental factors did not burden cardiovascular status. An initial total of 72 responses were obtained for demographic metrics and vital signs. Data on nicotine use and diabetes prevalence were compiled later, yielding 116 total responses.

#### Capacity Screening

Volunteers were instructed to proceed only if the individual was Alert and Oriented (A&O x4)—able to state their name, location, approximate time/day, and explain the purpose of the assessment (e.g., “You’re asking questions to help your organization”).

Exclusion Criteria with a “Hard Stop” protocol was implemented to ensure participants provided clear, voluntary, and informed consent and had the capacity to participate. The assessment was immediately discontinued if the individual appeared disoriented, confused, exhibited severely disorganized speech, appeared to be in a state of acute psychosis or severe paranoia, was under the acute influence of illicit drugs, or showed signs of severe intoxication that prevented understanding. Participants could withdraw consent at any time without penalty.

#### Eligibility Criteria

Strict eligibility criteria were applied to ensure the safety of all participants and to maintain the integrity and operational utility of the collected data. These criteria were designed to minimize potential risks, reduce variability introduced by confounding factors, and ensure that the study population appropriately represented the target group for which the findings are intended.

Inclusion criteria required participants to demonstrate sufficient capacity and orientation to engage meaningfully and provide informed consent in the survey. Participants were eligible for inclusion if they demonstrated alertness and orientation to person, place, time, and situation (A&O x4) with verbal consent obtained by the participant after any outstanding questions were answered.

The exclusion criteria were established to safeguard both participants and volunteers from potential harm and to ensure adherence to ethical standards. Individuals lacking decision-making capacity were excluded, as were those who withdrew or refused to proceed at any point in the encounter. Individuals displaying alterations in mental status or under the apparent influence of drugs or alcohol were excluded due to a lack of capacity or inability to provide informed consent. Stably housed individuals were also not eligible for participation. Figure 1 illustrates the screening process for participant eligibility, detailing the inclusion and exclusion criteria, capacity assessment, and consent procedures. This flow clarifies how participants were systematically evaluated to ensure ethical engagement, safety, and reliable data collection.

**Figure 1.**
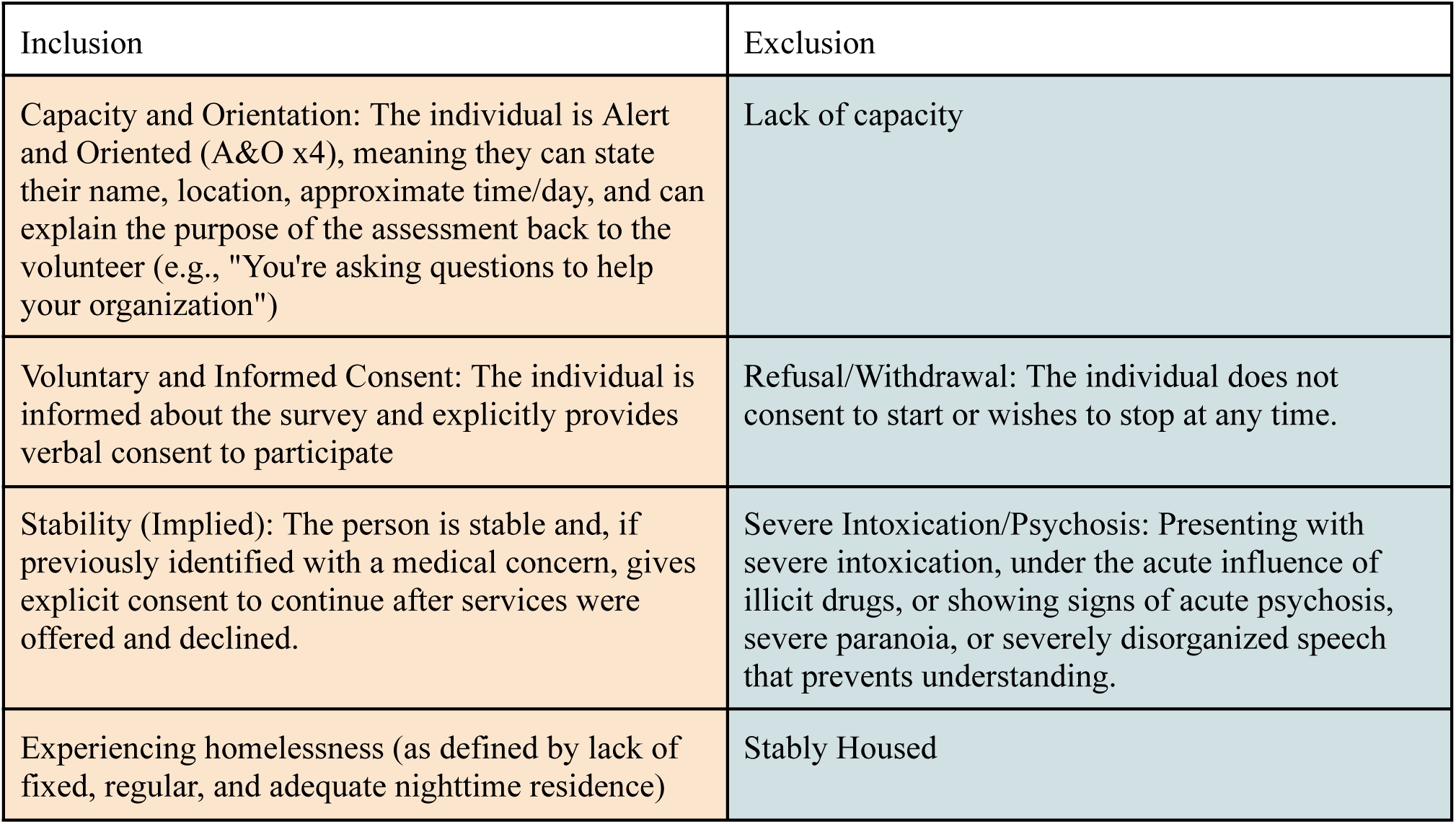
Flow of Participant Screening, Inclusion, and Exclusion Based on Eligibility Criteria.

#### Verbal Consent

Prior to data collection, volunteers read a Verbal Information & Consent Script confirming the activity was a needs assessment, not research. Participants were assured that participation was entirely voluntary, that they could stop at any time, and that their decision would not affect access to food or supplies. All answers were recorded anonymously with no names collected. Consent was obtained verbally after any questions were answered.

#### Data Collection

The questionnaire collected demographic data (age, gender, race/ethnicity), diabetes and nicotine use data, two physiological metrics, and one subjective metric: blood pressure, heart rate, and pain score, respectively.

Data was collected across the Inner Mission, Tenderloin, and Embarcadero neighborhoods of San Francisco, California. Outreach team members then approached participants and obtained informed consent, ensuring that participants appeared alert and oriented and had capacity.

Physiological measures were collected using FDA-cleared medical supplies, including automated wrist sphygmomanometers and finger pulse oximeters. BP monitoring devices include the G.Lab automatic blood pressure cuff and the Omron digital blood pressure cuff. The finger pulse oximeter devices used are CVS-brand devices. All devices were inspected for functionality to ensure the accuracy of measured vital signs and participants’ safety. Data were collected under the supervision of licensed medical professionals who oversaw the use of these devices and ensured proper use throughout the study.

In addition to physiological measures, subjective pain reporting was collected using the Numeric Pain Rating Scale (NPRS). The NPRS is utilized as a standardized tool to assess pain intensity. Participants were asked to rank their current pain levels on a scale of 1 to 10. Although standard practices used in clinical healthcare settings do include the ranking of “0” and participants may state a number greater than “10”, any rate provided as “0” will be transformed to “1” and any numerical value reported by a participant greater than 10 will be converted to “10” for data analysis purposes. To minimize literacy bias, surveys were administered orally to participants by members of the health team.

#### Emergency Response Protocol

The protocol prioritized participant well-being. Specific medical “red flags” were defined as abnormal vital sign readings that could indicate urgent or emergent clinical concern. This included elevated blood pressure (systolic pressure >180 or diastolic pressure >120) [17] and other metrics for abnormal heart rate, respiratory rate, and blood glucose readings. If a red flag was identified, the assessment was paused, and the participant was calmly informed of the potential danger. Immediate help was offered, such as calling 911. Volunteers documented vital signs and the actions taken (e.g., “BP 210/137. Offered to call 911; person declined”), and participants’ autonomy to decline services was respected.

If participants disclosed suicidal or homicidal ideation at any point during the survey, volunteers were trained to evaluate the level of risk and to dial 911 if the person was deemed to be an imminent risk to themselves or others within the next 24h. If a person is considered to have an A&O status of less than 4, volunteers are directed to activate the emergency response system by dialing 911.

### Statistical Methods Data Processing

All statistical analyses were performed in Microsoft Excel (Office 365), and some graphs were created in RStudio. Data were first screened for completeness and accuracy. Entries missing critical values were omitted from analysis, such as blood pressure measurements recorded as a single number (without a systolic/diastolic ratio). Pain scores reported as ranges were standardized by selecting the highest reported value (e.g., “6-7” was set to “7”). Any pain scores recorded as “0” were converted to “1” to align with the validated 1–10 pain scale. Likewise, any score reported to be above “10” was converted to “10”.

For descriptive statistics, the mean, median, and range were calculated for continuous variables, including age, heart rate, systolic blood pressure, diastolic blood pressure, and pain score. Based on established clinical thresholds, the frequency and percentage distributions were computed for categorical clinical parameters —specifically, heart rate and blood pressure categories.

For sex data, all entries containing “man” or “woman” were labeled as “male and “female” respectively. Additionally, entries with misspellings or abbreviations of male or female (e.g., Mal, M) were considered in the categories they abbreviated. For gender data, any explicit entry containing “transgender”, “non-binary”, or “gender nonconforming” was considered as its own separate category to ensure representation beyond the male and female binary. Sex is more determinant when considering the influences on biological risk factors for health outcomes, while gender is influential in social determinants of health and access to health care [18]. For race and ethnicity data, entries containing information from multiple categories (e.g., Irish & Latina, half Indian half White) were marked as “other/specific origin”. Entries containing single nationalities were also listed in the “other/specific origin” category unless the entry contained a nationality from a country of Spanish-speaking or Latin-American origin (e.g., Mexican), which were categorized as “Hispanic/Latino”. Entries not containing any data were labeled as “unknown/missing”.

Heart rates were categorized as: Bradycardia (<60 beats per minute [bpm]), Normal (60–100 bpm), and Tachycardia (>100 bpm).

Blood pressure was classified according to the American Heart Association (AHA) guidelines [19]: Normal: Systolic <120 mmHg and Diastolic <80 mmHg; Elevated: Systolic 120–129 mmHg and Diastolic <80 mmHg; Hypertension, Stage 1: Systolic 130–139 mmHg or Diastolic 80–89 mmHg; Hypertension, Stage 2: Systolic ≥140 mmHg or Diastolic ≥90 mmHg; Hypertensive Crisis (Emergency): Systolic >180 mmHg and/or Diastolic >120 mmHg; Hypotension: Systolic <90 mmHg or Diastolic <60 mmHg. Percentages of participants within each blood pressure and heart rate category were calculated relative to the total number of valid entries.

The Numerical Rating Scale (NRS) was used to measure pain intensity, a validated, widely used tool for pain assessment that can be administered verbally or visually via patient self-report [20]. The standard NRS ranges from 0 (“no pain”) to 10 (“worst pain imaginable”). To ensure analytic consistency in our dataset, all reported zeros were set to 1, resulting in a modified scale ranging from 1 to 10. Pain levels were categorized as mild (1–4), moderate (5–6), and severe (7–10). [20].

Participants disclosed diabetes data from 3 categories: type 1, type 2, and pre-diabetes. While there are notable commonalities and differences between type 1 diabetes mellitus (T1DM) and type 2 diabetes mellitus (T2DM), for analytical purposes, the diagnosis of “diabetes” includes both T1DM and T2DM. Both T1DM and T2DM, along with prediabetes, share mechanistic pathways for cardiovascular damage despite differing etiologies. To ensure analytic consistency and avoid statistical power or sample size concerns, given that T1DM is less common than T2DM, both T1DM, T2DM, and prediabetes were grouped as “diabetes” to create a more robust estimate of cardiovascular prevalence [21]. Participants who did not mention a history of diabetes when asked were, conversely, marked as “no diabetes” in the graphical data.

For nicotine usage data, any participant who was stated to have smoked, used cigars/cigarettes, or any other nicotine use, either at the time of survey or in the past, was counted in the “use nicotine” category. All other participants who did not report nicotine use were counted as “do not use nicotine”. Any diabetes or nicotine use data that participants did not report or surveyors could not record were excluded.

All results were summarized descriptively to guide operational health priorities rather than inferential conclusions. Graphical representations were generated using statistical visualization software to illustrate demographic distributions and clinical patterns. Given that this analysis was conducted as a community needs assessment and not human subjects research, no hypothesis testing, regression modeling, or inferential statistics were performed.

### Demographic Data

Of the initial 72 surveys, 70 provided quantifiable ages, which were used to calculate the Mean Age (42.96 years) and Median Age (41 years). The Age Range was 24 to 70.

#### Heart Rate (HR)

Analysis began with 72 total entries. Seven entries were excluded, including six non-numeric or unavailable entries (“na,” “NA”) and one physiological outlier exceeding the upper limit (825). The final set of 65 quantifiable entries (ranging from 55 to 130) was used to calculate the Mean Heart Rate (96) and Median Heart Rate (96).

#### Blood Pressure (BP)

Of the 72 initial Blood Pressure entries, 13 were omitted for missing/incomplete data, unclear notation, or values falling outside physiologically plausible ranges (e.g., 120/172, 135/32). The remaining 59 usable BP ratios were classified using standard AHA guidelines [22]

#### Self-Report Nicotine Usage

Analysis began with 116 total entries. Seventy-seven were excluded due to containing blank or “none reported/unable to record” responses. All entries that listed smoking or cigarette use were categorized as nicotine users alongside those who explicitly stated nicotine use. The resulting data yielded 39 usable entries.

#### Self-Reported Diabetes

Analysis began with 116 total entries. Seventy-eight entries were excluded due to containing blank or “none reported/unable to record” responses. All entries indicating any form of diabetes (Type 1 or Type 2) were grouped into a single “Diabetes” category for analysis. The resulting dataset included 38 usable entries.

## Results

Data from the community needs assessment on HR, BP, and pain levels yielded 72 responses, of which 70 were included in the demographic analysis after data cleaning. Physiological and subjective metrics yielded usable sample sizes ranging from N = 59 to N = 70 after applying predefined data-cleaning and transformation protocols of Nicotine and diabetes data were compiled at a later date. Of the 116 responses concerning nicotine use, 77 did not meet the inclusion criteria, resulting in 39 eligible responses for analysis. Similarly, of the 116 responses pertaining to diabetes prevalence, 38 met the inclusion criteria, while 78 were excluded.

### Demographics

Demographic information was collected from 72 total participants. Among the 68 responses where gender was explicitly defined, the cohort was predominantly male: 53 participants (73.61%) identified as male, 15 (20.83%) as female, and 4 (5.56%) had missing or undefined gender information. Figure 2 presents the gender distribution of participants in the community needs assessment, providing insight into the demographic composition of the unsheltered population surveyed.

**Figure 2.**
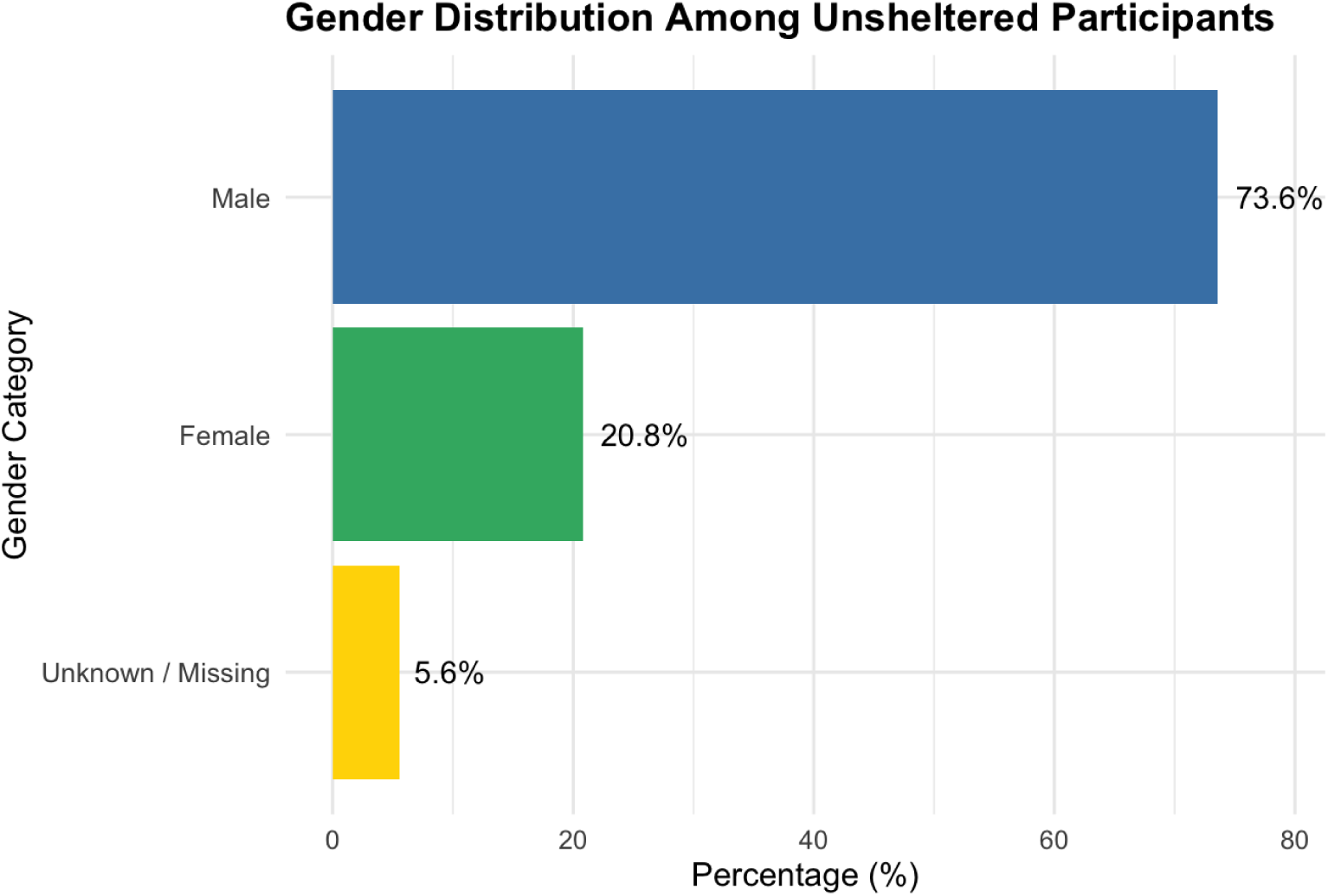
Gender Distribution Among Unsheltered Participants. The horizontal bar chart displays the gender composition of individuals in the community needs assessment. Among the 72 participants, 53 (73.61%) identified as male, 15 (20.83%) as female, and 4 (5.56%) had missing or undefined gender information.

### Age

A total of 70 responses provided quantifiable age data for analysis. Participant ages ranged from 24 to 70 years, with a mean age of 42.96 years and a median age of 41 years. Figure 3 illustrates the age distribution of participants in the community needs assessment, highlighting the demographic profile of the unsheltered population. Understanding age-related patterns is critical for tailoring health interventions, resource allocation, and outreach strategies to address the specific needs of different age groups within this vulnerable community.

**Figure 3.**
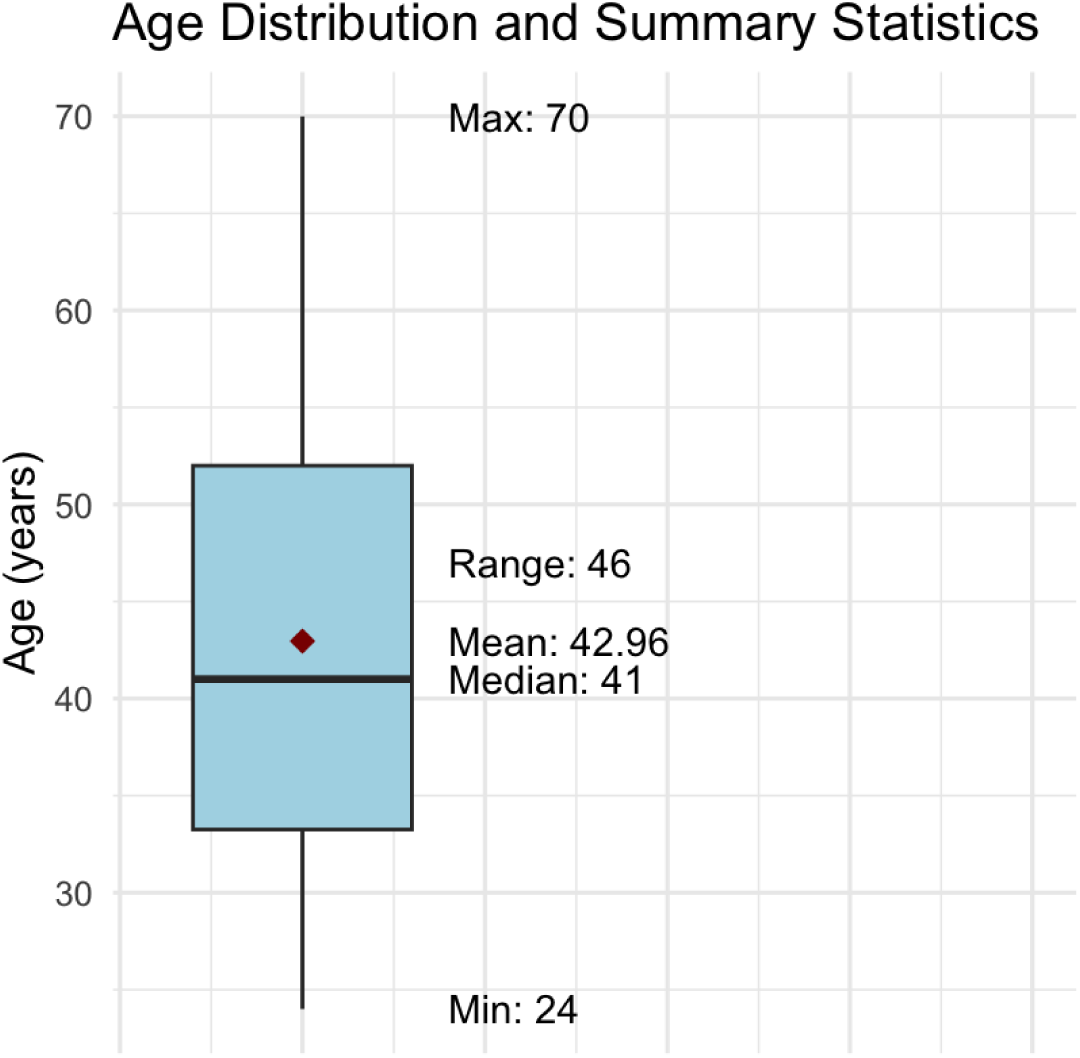
Age Distribution and Summary Statistics Among Unsheltered Participants. The boxplot summarizes the age distribution of unsheltered participants in the community needs assessment. Participant ages ranged from 24 to 70 years, with a mean age of 42.96 years and a median age of 41 years.

### Race and Ethnicity

Figure 4 illustrates the racial and ethnic composition of participants in the community needs assessment, providing critical context for understanding health disparities and informing culturally responsive interventions within the unsheltered population. This demographic information is important for identifying populations at higher risk, guiding culturally appropriate outreach, and ensuring that health interventions effectively address the specific needs of diverse racial and ethnic groups within the unsheltered community.

**Figure 4.**
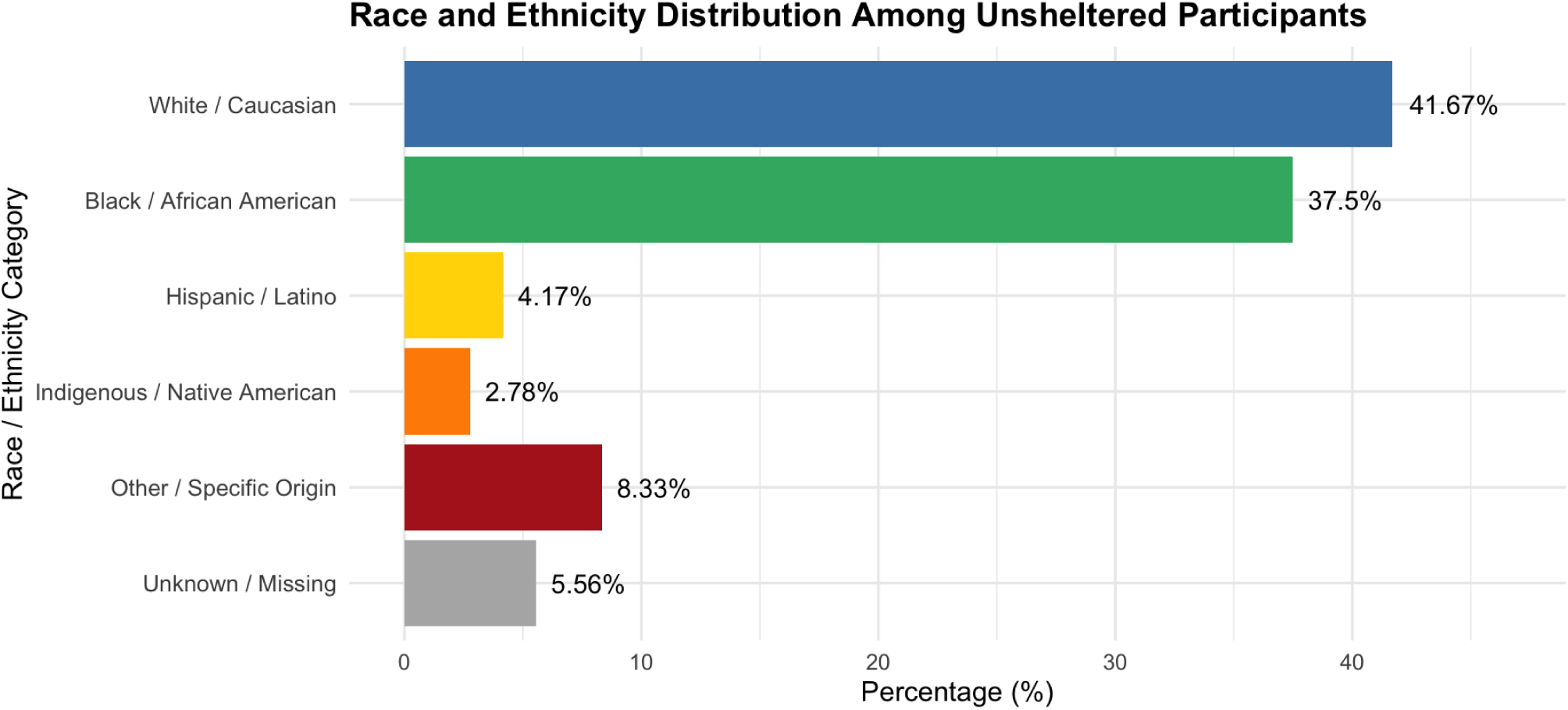
Race and Ethnicity Distribution Among Unsheltered Participants. Horizontal bar chart illustrating the distribution of racial and ethnic identities among unsheltered participants. The majority identified as White/Caucasian (41.67%) or Black/African American (37.50%), with smaller populations identifying as Hispanic/Latino (4.17%), Indigenous/Native American (2.78%), or Other/Specific Origin (8.33%). Data were missing for four participants (5.56%).

The cohort sample demonstrated racial and ethnic diversity. Of the 72 participants surveyed, the majority identified as White/Caucasian (N = 30) or Black/African American (N = 27). Smaller proportions of participants identified as Hispanic/Latino (N = 3), Indigenous/Native American (N = 2), or other/specific origins (N = 6). Data was missing or unavailable for 4 participants.

### Cardiovascular Metrics

Analysis of blood pressure began with 72 total data points. After excluding 10 non-reported, non-read, and ambiguous entries, alongside three physiological outliers (e.g., 120/172, 135/32), 59 usable BP ratios were retained for classification.

A substantial majority of participants presented with elevated or high blood pressure. Total High Blood Pressure (Hypertensive Stage 1 or Higher) accounted for 43 entries, representing 72.88% of the usable data points (N = 59).

Across the Usable BP Ratios (N = 59), the specific distribution across standard categories was as follows: Hypertensive Crisis or Higher: 6 entries (10.17%) met the threshold of SYS ≥180 OR DIA ≥120. These critical readings included 210/137, 286/127, 185/136, 193/140, 218/154, and 148/122. These findings triggered the Emergency Action Protocol outlined in the field guide. General Hypertensive (Stage 1 or 2): 37 entries (62.71%). Standard Range: 6 entries (10.17%). Elevated: 5 entries (8.47%). Hypotensive: 5 entries (8.47%).

Figure 5 illustrates the distribution of systolic blood pressure among participants, highlighting the range and central tendency of measurements after excluding invalid or physiologically implausible readings. This information is critical for identifying the prevalence of acute cardiovascular risk and informing targeted interventions, medical referrals, and emergency response planning within the unsheltered population.

**Figure 5.**
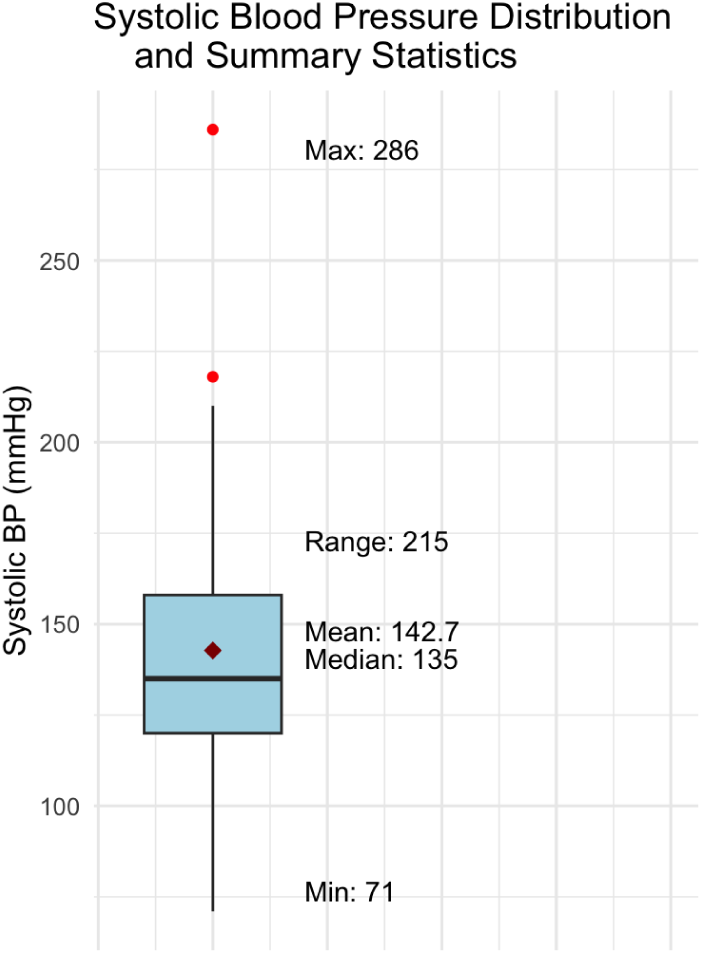
Systolic Blood Pressure Distribution and Summary Statistics Among Unsheltered Participants. The boxplot summarizes the distribution of systolic blood pressure among participants after excluding non-readings, ambiguous entries, and physiological outliers. Systolic blood pressure values ranged from 71 to 286 mmHg, with a mean of 143 mmHg and a median of 135 mmHg.

Figure 6 presents the distribution of diastolic blood pressure among participants after excluding non-readings, ambiguous entries, and physiologically implausible values. The observed range, mean, and median underscore elevated diastolic pressures in the cohort, underscoring the need for targeted cardiovascular risk management and timely clinical intervention in the unsheltered population.

**Figure 6.**
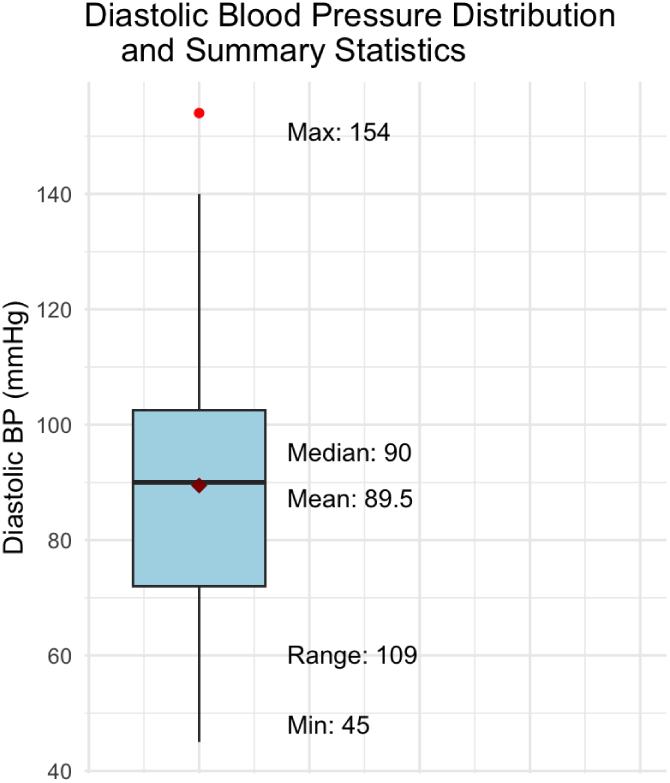
Diastolic Blood Pressure Distribution and Summary Statistics Among Unsheltered Participants. The boxplot summarizes the distribution of diastolic blood pressure among participants after excluding non-readings, ambiguous entries, and physiological outliers. Diastolic blood pressure values ranged from 45 to 154 mmHg, with a mean of 89 mmHg and a median of 90 mmHg.

Figure 7 illustrates the distribution of blood pressure categories among participants, as defined by the American Heart Association (AHA) guidelines. The data reveal a high prevalence of elevated and hypertensive readings, with nearly 57% of participants falling into Hypertension Stage 2 or Hypertensive Crisis, highlighting a critical need for urgent cardiovascular interventions, targeted health education, and structured referral pathways within the unsheltered population.

**Figure 7.**
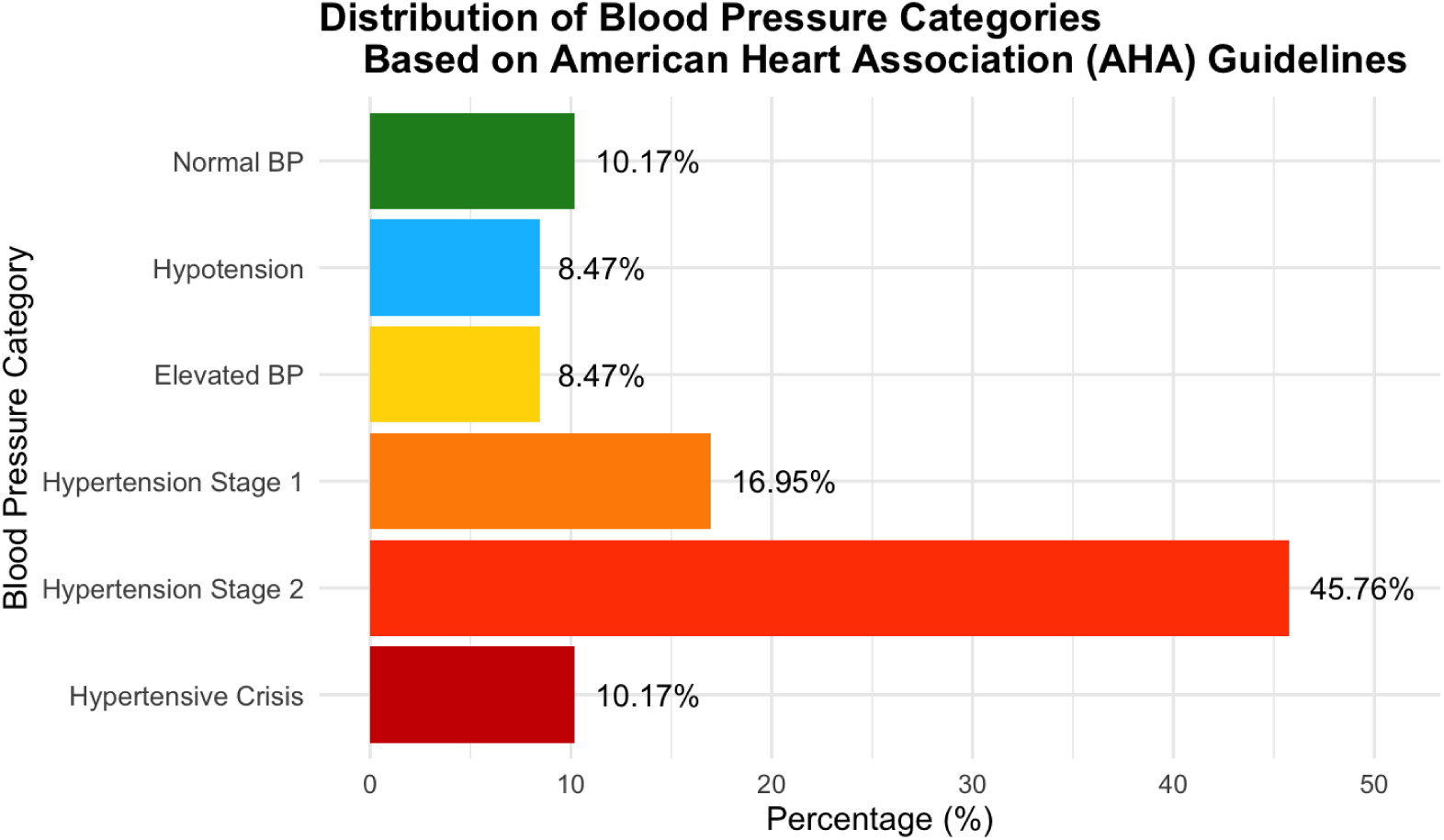
Distribution of Blood Pressure (BP) Categories Among Unsheltered Participants based on American Heart Association (AHA) Guidelines. Of the 72 total participants, 6 (10.17%) were classified as Normal, 5 (8.47%) as Hypotension, 5 (8.47%) as Elevated, 10 (16.95%) as Hypertension Stage 1, 27 (45.76%) as Hypertension Stage 2, and 6 (10.17%) as Hypertensive Crisis. Twelve participants had missing or unclassifiable readings.

Heart rate analysis, as shown in Figure 8, used 65 quantifiable entries from 72 original entries (ranging from 55 to 130), after seven exclusions (6 non-reported and one outlier above 200). The central tendency metrics indicate elevated baseline cardiac activity in the cohort. The Mean Heart Rate was 96, the Median Heart Rate was 96, and the range was found to be 55 to 130.

**Figure 8.**
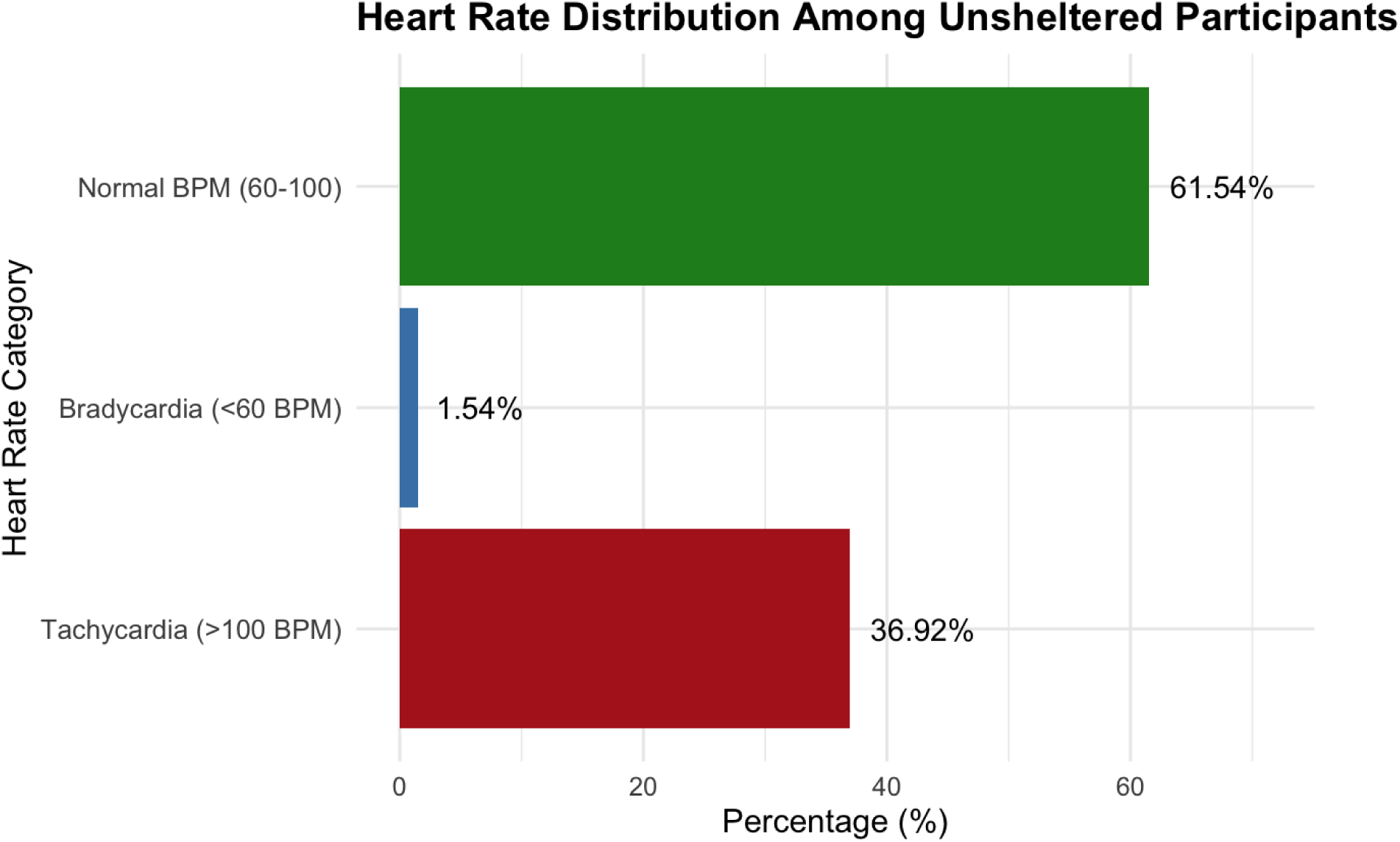
Distribution of Heart Rate Categories Among Unsheltered Participants. The horizontal bar graph illustrates the distribution of heart rate categories among unsheltered participants. From the data, 40 participants (61.54%) had heart rates within the normal range (60-100 bpm), 24 participants (36.92%) exhibited tachycardia (>100 bpm), and 1 participant (1.54%) exhibited bradycardia (<60 bpm). Heart rate data were missing or unclassifiable for 7 participants.

### Pain Score

Pain scale observations totaled 72 entries, yielding 66 usable scores after transformation and exclusions. The transformation rules applied were: 0→1 (seven entries transformed) and >10→10 (one entry transformed). The Numerical Pain Rating Scale was administered, with volunteers instructing participants that “1” meant no pain at all and “10” meant the worst pain they had ever felt. Pain Score Metrics (N = 66). The Mean Pain Score was 3.74, the Median Pain Score was 3.0, and the Pain Score Range was 1 to 10.

Chronic symptom burden was significant, with a substantial percentage reporting severe pain. Scores of 1 accounted for 23 entries (34.85%). Meanwhile, scores of 2-6 accounted for 30 entries (45.45%). Scores between 7 and 10 (Severe Pain) accounted for 13 entries (19.70%). Figure 9 presents the distribution of pain scores among unsheltered participants, offering a visual summary of the varying pain levels reported within this population. This figure allows for a clearer understanding of the prevalence and intensity of pain experiences, emphasizing the need for accessible pain management and supportive care.

**Figure 9.**
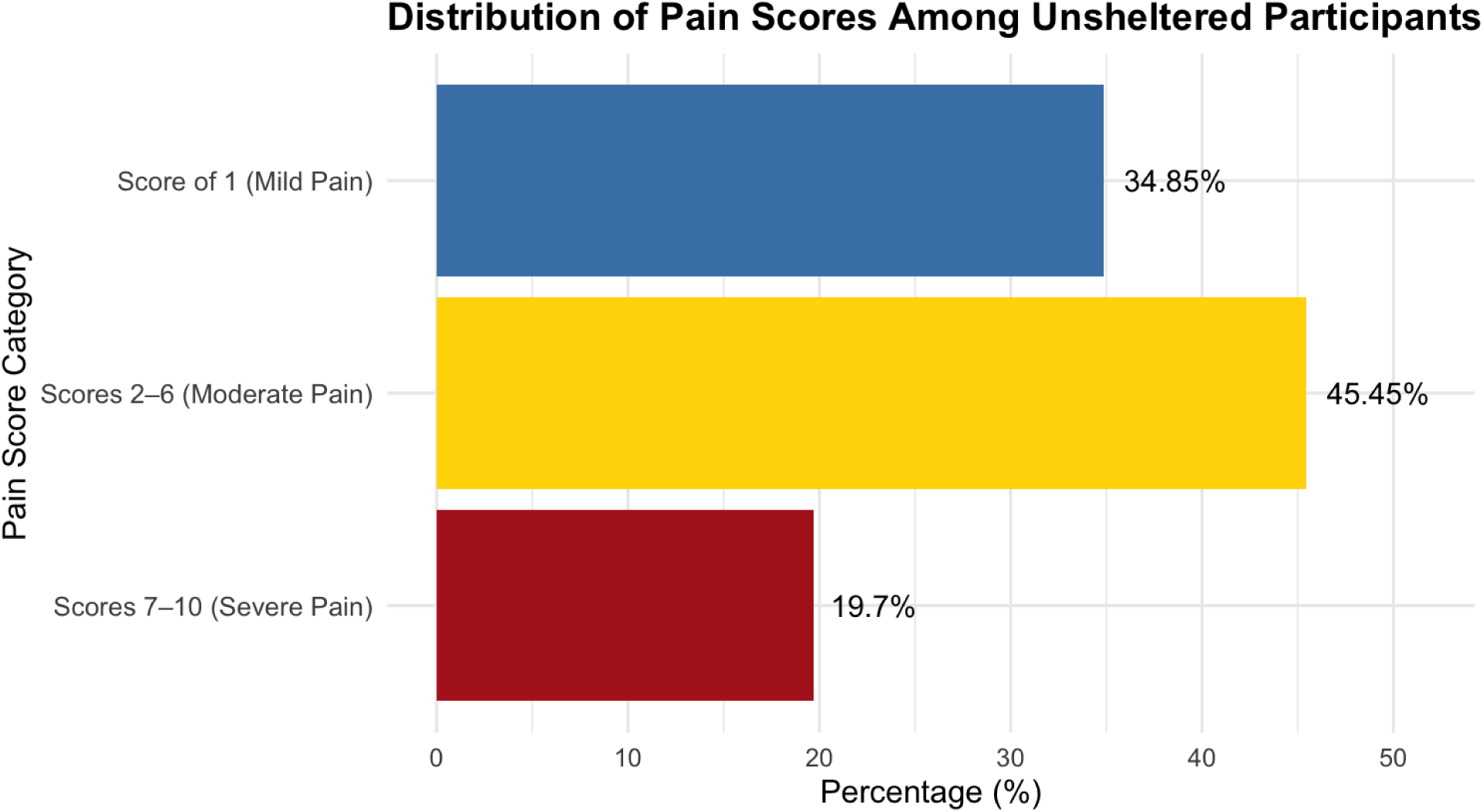
Distribution of Pain Scores Among Unsheltered Participants. The horizontal bar graph illustrates the distribution of pain scores among unsheltered participants. From the data, 23 participants (34.85%) had a mild pain score of 1, 30 participants (45.45%) had a moderate pain score of 2-6, and 13 participants (19.70%) had a severe pain score of 7-10. Pain score data were missing or unclassifiable for 6 participants.

### Diabetes

All health data were collected through standardized clinical and self-report questionnaires administered by trained outreach volunteers. The objective metrics, including heart rate (HR) and blood pressure (BP were obtained through point-of-care measurements. Among participants, 5 (13.16%) reported a history of type 1 diabetes, type 2 diabetes, or pre-diabetes; 33 (86.84%) reported no history of diabetes. Figure 10 illustrates the distribution of diabetes among unsheltered participants, highlighting the proportion of individuals affected by this chronic condition.

**Figure 10.**
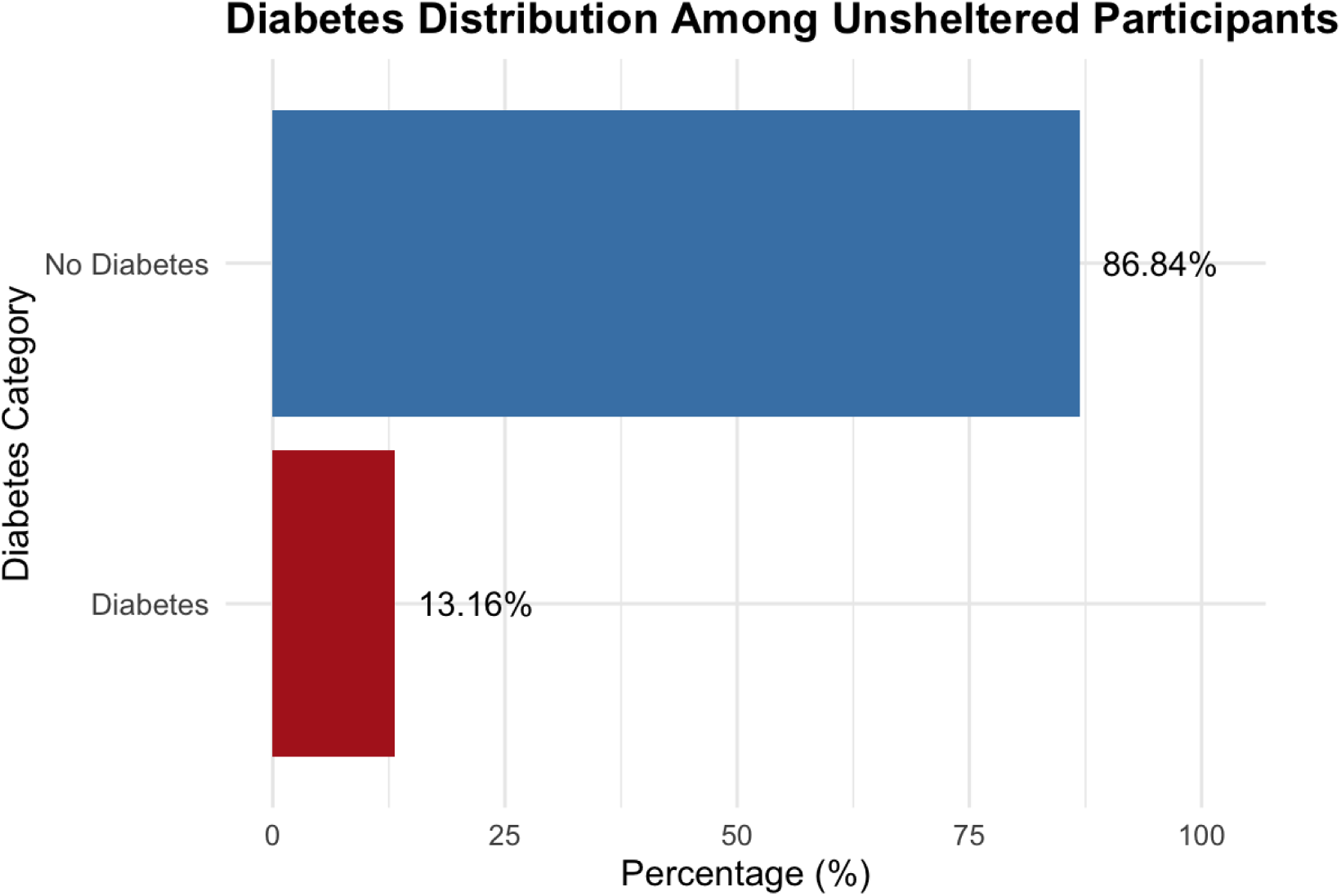
Distribution of Diabetes Among Unsheltered Participants. This horizontal bar graph shows the prevalence of diabetes among unsheltered participants. Of the total participants, 33 (86.84%) reported no diabetes, and 5 (13.16%) had diabetes.

### Nicotine Use

Information on nicotine use was collected through a structured self-report questionnaire administered during field outreach. Participants were asked whether they currently use nicotine products. Figure 11 presents the distribution of self-reported nicotine usage. The data consisted of 116 entries, of which 39 were usable. Among the participants, 33 (84.62%) reported no nicotine use, whereas 6 (15.38%) reported current nicotine use.

**Figure 11.**
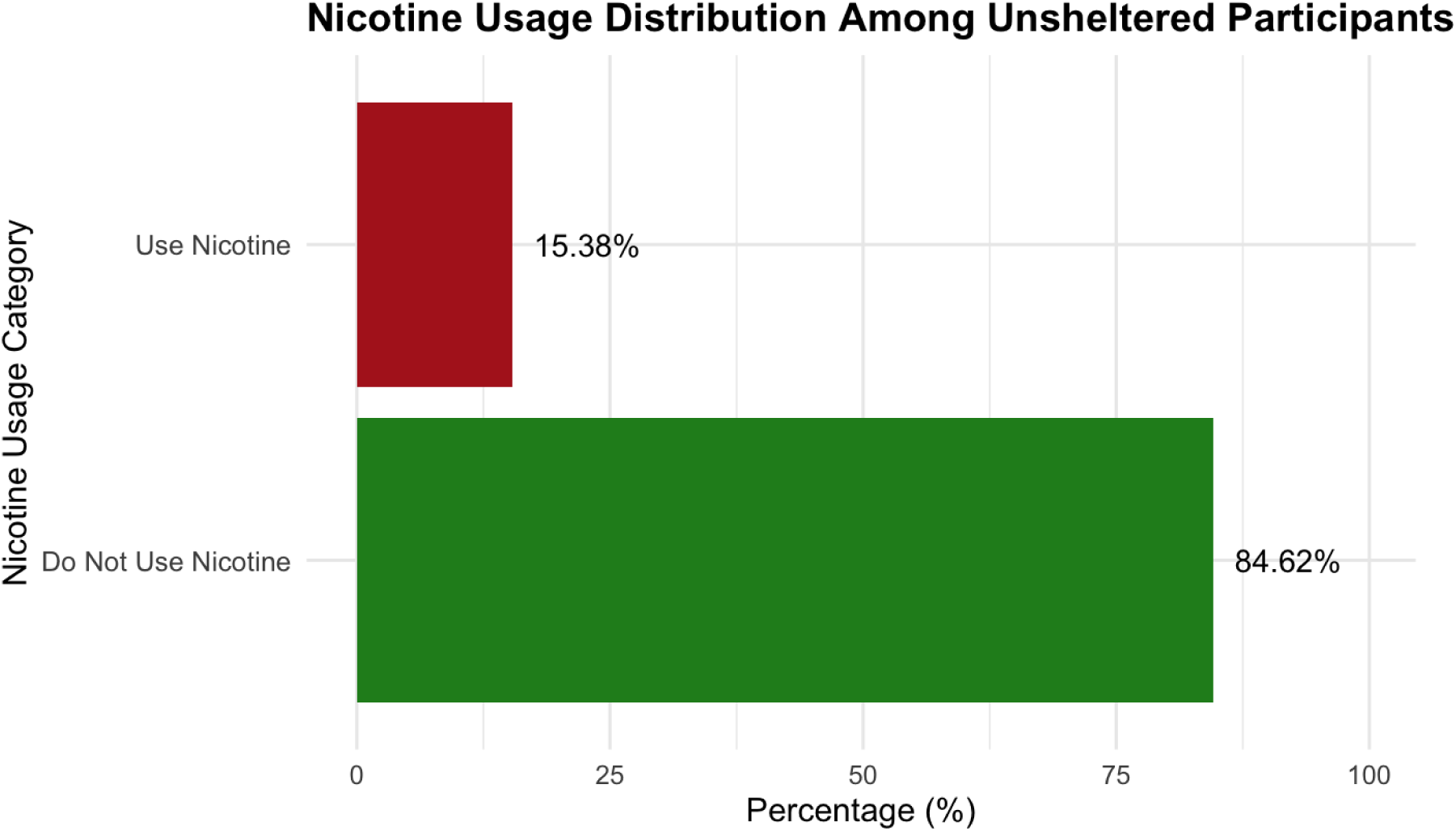
Distribution of Nicotine Usage Among Unsheltered Participants. This horizontal bar graph illustrates nicotine usage among the unsheltered participants. Of the participants, 33 (84.62%) reported no nicotine use, and 6 (15.38%) reported nicotine use.

## Discussion

This descriptive public health report details the demographic and clinical characteristics of an unsheltered population, providing quantitative evidence of significant acute health crises that are often unmanaged. Framed strictly as an Operational Needs Assessment (ONA), the findings serve to justify the urgent expansion of health outreach services, health education, and partnerships.

### Ethical Framing and Operational Necessity

A crucial element of this report is its foundation as a community needs assessment rather than a research study. This approach was overseen by an independent ethics committee, which determined the status of NHSR. This ethical framework mandated a stringent “Hard Stop” protocol that prioritized participant well-being, informed consent capacity, and data collection autonomy. The primary utility of the data, therefore, is its direct application to the operational decisions of Valliant Foundation, guiding the prioritization of referrals, educational materials, and services. This report fulfills the operational mandate to better understand community needs and justify resource allocation by detailing the prevalence of critical health concerns.

### The Prevalence of Acute Cardiovascular Risk

The most compelling finding is the extremely high prevalence of severe cardiovascular risk factors, which warrants immediate clinical attention. Of the 59 usable BP ratios, a staggering 72.88% (N=43) were classified as Total High Blood Pressure (Hypertensive Stage 1 or higher). This suggests that hypertension is not a statistical anomaly but a dominant, pervasive medical condition within this cohort. The data were benchmarked against national and published rates to help determine baseline ranges for the clinical values. According to recent national data, the general American adult population reports that hypertension prevalence among adults 18 and older is about 47.7%, providing a baseline comparison to our cohort data. This varies across age groups and genders, ranging from 34.6-59.2% when examining the total of adults 18 and over and their awareness of hypertension status. [23].

In contrast, the rate in this assessment, 72.88% total high blood pressure and 10.17% in hypertensive crisis, is markedly higher. Elevated cardiac activity supports this interpretation: the Mean Heart Rate of 96 bpm and 36.92% tachycardia rate significantly exceed expected norms (18–22%), suggesting chronic sympathetic activation and heightened stress response.

More alarmingly, 10.17% (N=6) of the usable readings met the criteria for Hypertensive Crisis or Higher (SYS ≥180 OR DIA ≥120). These critical measurements, which included values up to 286/127 and 218/154, necessitated implementing the Emergency Action Protocol outlined in the field guide, which informed participants of the danger and offered immediate help, such as calling 911. The high rate of these extreme outliers strongly suggests that many unsheltered individuals are living with severe, untreated, or poorly managed hypertension, putting them at extreme risk for myocardial infarction, stroke, and kidney failure.

Furthermore, heart rate assessment revealed elevated baseline cardiac activity. The Mean Heart Rate (96) and Median Heart Rate (96) (N=65), with a maximum recorded rate of 130, are above healthy resting averages. This may be indicative of chronic physiological stress, anxiety disorders, or unmanaged pain, all of which are common comorbidities in the unsheltered population. Another consideration for elevation in heart rate would be the use of nicotine or tobacco products, given that nicotine/tobacco acutely raises HR and BP. Previous studies reported approximately +11.3 bpm in HR and +12.9 mmHg in SBP, +7.7 mmHg in DBP [24] among nicotine users, underscoring that increases in cardiovascular metrics may not be limited to environmental and physiological stressors but may also be driven by nicotine and tobacco product consumption. The combined findings of severe hypertension and elevated heart rate present a composite picture of imminent cardiovascular danger that health outreach programs must urgently address.

### Chronic Symptom Burden and Pain

Pain scores were subjective values reported by participants using the Numeric Pain Rating Scale (NPRS; 1-10) to assess pain at the moment of the encounter between the participant and the volunteers collecting the survey. A transformation was applied upon reviewing the data to ensure the accuracy of the descriptive statistics. The transformation protocol required that any pain score reported as less than 1, such as “0”, be transformed to “1”, and any score >10 be converted to “10”.

Applying this adjustment operationalized a score of 1 as the benchmark for “no pain at all,” as any value below one would distort the true prevalence of other reported pain values. Additionally, several entries illustrated the complexity of pain assessment in the field. For example, one participant who had recently been struck by a motorcycle reported a current pain score of six. This highlights the difficulty in distinguishing acute trauma from chronic pain, reinforcing the need for careful, ethically informed data collection. Values such as this one were not transformed, but it is worth noting that some values may be underrated due to feelings of burdensomeness or desensitization.

After transformations were applied and descriptive statistics were employed, a Mean Pain Score of 3.74 (N=65) was calculated. Notably, nearly one-quarter of the participants (19.70%, N=13) reported severe pain scores between 7 and 10. This high rate of severe pain underscores the chronic physical distress experienced by this population, which demands comprehensive pain management and referral services. Pain is a potential driver of elevated heart rate and complicates the management of underlying conditions, providing a crucial intervention point for improving quality of life and cardiovascular health.

### Demographic Context

The demographic profile reveals a predominantly male cohort (55 entries) and a large concentration of middle-aged adults (Mean Age 42.96 years), consistent with many national reports on unsheltered populations. The near-even split between White/Caucasian (30 entries) and Black/African American (27 entries) participants indicates the urgent necessity of developing culturally competent and equitable health interventions that address the unique needs and barriers faced by these two largest demographic groups. For example, in-field culturally competent health education could include peer-led blood pressure workshops facilitated by trained community health workers from similar racial or cultural backgrounds. They can use accessible language and relevant examples to explain hypertension management, medication adherence, and nutrition within local resources. Such approaches have been shown to build trust, improve engagement, and enhance understanding among African-American participants.

Though demographic data collected demonstrates a comparable number of Black/African American and White identifying participants that are predominantly middle-aged and male, this reflects the intersection between race, gender, and health vulnerability. Public health data from San Francisco in 2024 revealed that 37% of unhoused adults identified as Black while only 5% of San Francisco residents are Black [25] Data from this study supports overrepresentation of Black/African Americans experiencing homelessness in San Francisco established by existing demographic data about the unhoused population of San Francisco [26] The high rates of untreated or inadequately managed hypertension observed in this study reflect a barrier that is unique to the unsheltered adult community as they often lack access to preventative care measures and treatment. Other considerations for overrepresentation of unhoused Black/African Americans when observing cardiovascular risks can be attributable to the history of fear and mistrust among Black/African Americans and the healthcare system, as it has been noted that stress levels are correlated to levels of fear and mistrust in healthcare, and these sentiments have been found to increase under unsheltered circumstances [27]

### Recommendations For Program Improvement

Consequently, the findings also prompt immediate, practical action. We strongly recommend increasing referrals to cardiovascular clinics for homeless individuals at higher risk, conducting further NHSR analysis, or possibly future hypothesis-driven research to better understand health risks within this population. These steps ensure improved preventative care and more effective future interventions.

Given the disproportionate amount of Black/African American participants facing burdensome BP and HR values, it is essential to prioritize equity-centered approaches when considering interventions. This includes creating community engagement in outreach, training volunteers to understand biases and cultural humility to avoid dismissal of symptoms, and on a grander scale, proposing structural reforms that address systemic racism in housing and healthcare [28]

Further, the high prevalence of hypertension suggests possible overlap with undiagnosed or poorly managed diabetes, as both conditions frequently coexist in medically underserved populations. Incorporating blood glucose screening, diabetes education, and nutrition counseling during outreach events could enhance early detection and prevention. Similarly, because tobacco use significantly contributes to elevated BP and HR, implementing smoking cessation initiatives, including education, nicotine replacement guides, and emotional support, would further reduce cardiovascular risk and improve long-term outcomes.

The documented high rates of Hypertensive Crisis (10.17%) and elevated heart rates establish a compelling rationale for subsequent, rigorous meta-analyses to investigate associations between housing status, chronic stress, pain burden, and cardiovascular outcomes in this community. The success in collecting and cleaning multi-metric data (with high usability rates for BP, HR, and pain scores) demonstrates the feasibility of executing other protocols in a street-based setting. The methodology holds potential for other outreach service providers to emulate and implement their own program evaluation.

Valliant Foundation translated these findings into concrete operational changes to boost emergency training protocols and field education. The 10.17% rate of BP readings classified as a hypertensive crisis urges the creation of dedicated emergency referrals and transportation funding to ensure immediate pathways to emergent medical care for critical cases when unhoused persons decline EMS activation. To maintain best practices aligned with clinical protocols, it is advisable to periodically retrain volunteers to recognize hypertensive risk profiles and promptly initiate referrals during outreach. In addition, Valliant Foundation should incorporate targeted education for participants on diabetes management, including recognizing symptoms of hyperglycemia and hypoglycemia, understanding the importance of regular glucose monitoring, and connecting individuals to appropriate care, as well as smoking cessation, including providing brief interventions, distributing educational materials, and offering referrals to cessation programs or support services. Such suggestions reinforce the idea that the NHSR findings are descriptive and can be translated directly into practices and procedures to improve client safety and health outcomes.

### Limitations

As an operational needs assessment, this report is subject to limitations inherent to field collection. It was conducted as a community needs assessment rather than a formal research study, meaning that data were gathered solely to guide operational decision-making and resource allocation for outreach activities within a specific unsheltered population. As such, the data are descriptive and non-inferential, designed to identify urgent health priorities rather than to generate statistically generalizable findings. These results should not be interpreted as representative of the broader homeless population or as generalizable outcomes. The sample reflects individuals encountered during field operations within a defined geographic area and timeframe and is inherently limited by selection bias and situational factors.

Additionally, the assessment relied on self-report questionnaires for several health and behavioral indicators. Self-reported data are subject to recall bias, social desirability bias, and misreporting, which can lead to underestimation or overestimation of certain health conditions and behaviors. Unlike objective clinical measures, self-report instruments may not capture the full extent of undiagnosed or unmanaged conditions, limiting some findings’ accuracy and reliability. These findings provide a clear snapshot of the community’s immediate health needs.

The exclusion of nine BP entries (due to non-reads or physiological improbability) and six HR entries (non-reads or outliers) suggests inherent challenges in capturing precise physiological data in a non-clinical environment. However, the severity and volume of the remaining quantifiable high-risk findings (e.g., N=6 in Hypertensive Crisis) remain highly persuasive and mandate urgent operational response.

Participation required individuals to be alert and oriented; therefore, those experiencing severe confusion, acute psychosis, or intoxication were excluded under the protocol. This necessary safeguard introduces potential selection bias and likely underestimates the actual severity of the health crisis, as individuals with the most significant clinical instability were ethically ineligible to participate.

## Conclusion

This document presents the synthesized results of an ongoing Operational Needs Assessment (ONA) conducted by Valliant Foundation, which an independent ethics committee determined to be Non-Human Subjects Research (NHSR). The descriptive analysis of the cohort (N=116 total responses) establishes clear, urgent health priorities and confirms an “acute and ongoing ‘silent emergency’” within the unsheltered community, necessitating immediate operational strategic planning.

The quantitative findings reveal a profound and unmanaged cardiovascular crisis. A critical 10.17% (N=6) of the 59 usable BP readings met the threshold for Hypertensive Crisis or Higher (SYS ≥180 or DIA ≥120). This extreme rate, underscored by documented critical values up to 286/127 and 218/154, validates the ethical requirement for the Emergency Action Protocol. Furthermore, most of the cohort, 72.88% (N=43) of usable BP readings, were classified as Total High Blood Pressure (Stage 1 or higher). This prevalence is markedly higher than published national and unsheltered norms, underscoring the pervasive nature of severe hypertension within this specific population.

Elevated cardiac activity (Mean Heart Rate 96 bpm) and high rates of Severe Pain (19.70%) serve as crucial indicators of chronic physiological stress and comorbidities that complicate underlying cardiovascular risk. Tobacco use emerged as a significant health behavior, with high prevalence among participants, further compounding cardiovascular risk and complicating long-term disease management. Integrating targeted smoking cessation education and referral pathways into outreach missions is essential to address this modifiable risk factor. Additionally, self-reported diabetes prevalence highlights gaps in disease monitoring or access to consistent care, underscoring the need for targeted diabetes education, screening, and linkage to services.

Culturally competent health education is particularly essential for effectively addressing hypertension disparities among African-American populations, who experience disproportionately higher rates of uncontrolled blood pressure and cardiovascular complications. Historical patterns of fear and mistrust toward the healthcare system among Black/African-Americans, rooted in systemic racism and unethical medical practices, continue to influence engagement and access to care. Interventions such as Faith-based community programs, blood pressure self-management workshops, and the use of trusted community health workers have demonstrated improved engagement, adherence to medication, and lifestyle modification among African-American groups. Integrating similar culturally responsive approaches into Outreach efforts can enhance trust, communication, and overall intervention efficacy within diverse unsheltered populations.

This operational assessment underscores the imperative for Valliant Foundation to reassess its service delivery model in response to the acute and chronic health burdens identified among unsheltered individuals experiencing homelessness. The data necessitate an ethical obligation to allocate substantial resources toward establishing urgent medical referral pathways, providing transportation assistance for participants with blood pressure readings within the Hypertensive Crisis range or severe hypertension, and developing culturally competent outreach programs.

For instance, the National Hypertension Control Initiative (NHCI) demonstrated that community health centers employing self-measured blood pressure interventions, coupled with culturally sensitive education, achieved a 12.3% increase in blood pressure control rates from 2020 to 2022 among at-risk populations, including Black, Hispanic, and American Indian/Alaskan Native communities [29]. This approach aligns with evidence supporting integrating culturally appropriate hypertension education and community-based interventions to enhance medication adherence and lifestyle modifications [30].

Additionally, implementing team-based care strategies, as the Community Preventive Services Task Force recommends, has improved blood pressure control across diverse populations. These strategies encompass patient follow-up, medication management, self-management support, and self-measured blood pressure devices, all enhancing patient engagement and adherence to treatment protocols [31].

These findings underscore the complex intersection of social determinants, historical trauma, and healthcare inequities contributing to cardiovascular risk. Addressing these disparities requires culturally informed, trust-building interventions prioritizing continuity of care, psychological safety, and equitable access to preventive cardiovascular services within unsheltered and marginalized communities.

Ultimately, the NHSR initiative achieved its strategic objectives by generating robust, multi-metric data while upholding participant autonomy, informed consent, and well-being. The findings demonstrate the feasibility of implementing rigorous protocols in street-based settings and provide essential metrics to support partnership development. This evidence can guide future operational decisions to optimize participant health and safety.

## Data Availability

All data produced in the present work are contained in the manuscript

## Other Information

## Declaration of generative AI and AI-assisted technologies in the manuscript preparation process

During the preparation of this work, the authors used NotebookLM to improve grammatical, spelling, and organizational clarity. After using this service, the authors reviewed and edited the content as needed and took full responsibility for the published article.

## Acknowledgments

The authors are grateful for the operational and ethical support integral to this community needs assessment, which was determined as Non-Human Subjects Research (NHSR).

We thank Valliant Foundation for providing all necessary medical equipment and dedicated personnel to enable the collection of field data. We specifically acknowledge the meticulous work of the field data collection team, who successfully implemented the “Hard Stop” Protocol and the Emergency Action Protocol: Yacoob Modan, Kelly Tang, Ileana Jade Rodriguez, Sarah Jean Valliant, Angelina Ka Lee, Macarena Sofia Puig, Roxanne Carlos Mendoza, Raquel Juliette Tamayo, Trevor Anderson, Logan Holbrook, Sean M. Velasquez, Natalia Moonsinghe, Tyla Williams, Destiny-Elizabeth Silva-Castro, Rianna Punzalan, Samantha Parton, Elvin Sanchez Segundo, Zaneb Mehmood, Nayab Mehmood, Simran Athwa, Melanie Paulette Magallanes, Melissa Hernandez-Monroy, Belle Botelho, Katherine Mejia, Jesus Ernesto Salcedo, Jacob R. Derin, Oishee Maharatna, Andrew Monterola, Abigail Restivo, Crow Nguyen, Maya Barbosa, Jocelyn Garcia, Ashley Zhao, Tiffany, and Manuel Medina Estrada. We would also like to thank our Chief Operating Officer Kelly Tang for coordinating the data collection missions and overseeing the training of new volunteers. The authors also thank Aaron Sullivan for his valuable feedback, including his recommendation to incorporate smoking and diabetes as key cardiovascular risk factors within the study framework.

## Author Contributions

S.J.V. conceived and led the project as principal investigator and corresponding author, oversaw methodological design, ethical and regulatory compliance, data analysis, funding acquisition, and manuscript writing. I.J.R. served as project manager, coordinating data collection and contributing to the interpretation of physiologic metrics and manuscript drafting. C.M.K. served as the faculty advisor, guiding all phases of the project. C.M.K. reviewed and collaborated with S.J.V. on the study design, analysis, and final manuscript to ensure academic rigor and integrity. A.K.L. contributed to the writing of the manuscript, literature review, and citation management. K.T. was the Chief Operating Officer and oversaw data collection missions and the emergency action protocol. L.C.H., R.R.P., aided in statistical analysis of the data and oversaw the design of figures. R.J.T., R.C.M., M.S.P., T.A., Y.E.M., S.A., I.L., M.H.M., D.E.A.S.C., and all additional listed contributors assisted in data collection, participant engagement, and manuscript review. All authors approved the final version of this report.

